# Uncertainty-aware prediction of 48-month eGFR decline in type 2 diabetes mellitus: a secondary analysis of ACCORD

**DOI:** 10.64898/2026.08.04.26359704

**Authors:** Daniel Olshvang, Carl Harris, Rama Chellappa, Chirag Parikh, Prasanna Santhanam

## Abstract

**Background:** Long-horizon kidney trajectory prediction in type 2 diabetes mellitus (T2DM) is usually reported as a point estimate or event risk, although clinical decision-making also depends on whether an individual prediction is reliable. We developed an uncertainty-aware model for 48-month estimated glomerular filtration rate (eGFR) decline and tested whether conformal interval width provides a clinically structured, patient-level signal of prediction reliability.

**Methods:** We performed a secondary prognostic modeling analysis of Action to Control Cardiovascular Risk in Diabetes (ACCORD) participants with baseline and 48-month eGFR *(n=*6,853). The outcome was annualized eGFR change, calculated as 48-month minus baseline eGFR divided by four years. The primary baseline feature set excluded serum creatinine since eGFR is creatinine-derived, and also excluded urine biomarkers. Random forest, gradient boosting, penalized linear models, and XGBoost were compared using fixed training, calibration, and test partitions. Split and locally adaptive conformal intervals were evaluated by empirical coverage and interval width. Interval-width analyses were repeated after conditioning on baseline eGFR.

**Results:** The best primary model was random forest (R^2^=0.382, MAE=3.271 mL/min/1.73m^2^). Split 90% conformal intervals achieved empirical coverage of 0.917. Locally adaptive 90% intervals achieved empirical coverage of 0.909 with mean width 13.759 mL/min/1.73m^2^. In unadjusted analyses, wider intervals were associated with larger errors and more rapid decline. After interval-width quintiles were assigned within baseline-eGFR strata, wider intervals remained associated with realized prediction error (annual adjusted increase, 0.151 mL/min/1.73m^2^ per quintile). Beyond baseline eGFR, wider intervals were associated with younger age, female sex, higher HbA1c, higher triglycerides, and higher systolic blood pressure.

**Conclusions:** Baseline clinical variables predicted 48-month eGFR decline with good long-horizon performance in ACCORD, even after excluding serum creatinine and urine biomarkers from the primary model. Conformal prediction provided calibrated patient-specific intervals, and interval width behaved as an informative reliability phenotype rather than a random modeling artifact. These findings support a novel uncertainty-aware framing of kidney trajectory prediction in which rapid and uncertain decline can be identified from baseline clinical data.

## Background

Diabetic kidney disease is usually monitored through repeated eGFR and urine albumin measurements, which contemporary chronic kidney disease guidance treats as central markers for staging, monitoring, and risk assessment [1]. Diabetes-specific kidney guidance similarly emphasizes serial kidney-function and albuminuria assessment as part of longitudinal risk management, as The American Diabetes Association recommends regular chronic kidney disease (CKD) screening and risk-based management in diabetes care [2]. For patients and clinicians, the practical question is often not only whether kidney failure will eventually occur, but how quickly kidney function is likely to change over a clinically meaningful horizon [3]. Annualized eGFR change over 48 months is therefore attractive because it is interpretable, continuous, and aligned with longitudinal clinical monitoring [4].

Existing kidney risk models have had major clinical impact, particularly for predicting kidney failure in established chronic kidney disease [5]. However, models for medium-term eGFR trajectory in adults with type 2 diabetes mellitus (T2DM) are less established. In addition, many models emphasize point prediction while giving less attention to patient-specific uncertainty. Medical machine-learning guidance increasingly emphasizes that prediction tools should support clinical decisions by communicating both expected risk and the reliability of model output [6,7]. A predicted decline of 3 mL/min/1.73 m^2^ per year should be interpreted differently when the uncertainty interval is narrow versus very wide. A narrow interval suggests that the model’s estimate is relatively reliable for that individual, whereas a wide interval indicates that clinically important alternative trajectories remain plausible despite the same point prediction.

Conformal prediction provides a model-agnostic framework for calibrated prediction sets and intervals [8,9]. Distribution-free conformal inference has been developed for regression settings in which valid predictive intervals are needed around continuous outcomes [10]. Conformalized quantile regression and related approaches further show how interval width can adapt to heterogeneity in residual variability [11]. Recent medical and machine-learning tutorials have emphasized conformal prediction as a practical route to uncertainty-aware decision support [12]. This is important for medical informatics because the model output becomes a paired estimate: the expected trajectory and the reliability of that trajectory for an individual patient.

We used the Action to Control Cardiovascular Risk in Diabetes (ACCORD) trial, a large multicenter randomized study of adults with T2DM at high cardiovascular risk, to develop and validate an uncertainty-aware model for 48-month eGFR decline [13]. ACCORD was a multicenter North American randomized trial that included more than 10,000 adults aged 40 to 79 years with T2DM and either established cardiovascular disease or multiple cardiovascular risk factors. Participants were assigned to intensive or standard approaches to glycemic control, with additional randomization to blood pressure and lipid interventions in eligible subgroups. The parent glycemia trial provides the core randomized-treatment structure and long-term metabolic-risk context [14]. The embedded blood-pressure trial adds a second randomized cardiovascular-risk intervention relevant to kidney trajectory modeling [15]. The embedded lipid trial contributes fenofibrate assignment and lipid phenotyping, both of which are relevant to diabetic microvascular risk [16]. ACCORD microvascular analyses established kidney-related outcomes as clinically important trial endpoints in this population [17]. Our primary goals were to determine whether baseline clinical variables could predict long-horizon eGFR change, and to test whether conformal interval width identifies a clinically meaningful phenotype of rapid and uncertain kidney decline.

## Methods

### Study population

We conducted a secondary analysis of ACCORD, a multicenter randomized trial that enrolled adults with T2DM at high cardiovascular risk. For the present analysis, the study cohort was restricted to participants with both baseline and 48-month eGFR measurements available, permitting calculation of annualized 48-month eGFR change. Exclusions were due to unavailable baseline eGFR (n=56), death before 48 months (n=471), severe renal outcome before 48 months (n=59), recorded follow-up ending before 48 months (n=1,591), and unavailable 48-month eGFR despite follow-up through 48 months (n=1,221). Baseline characteristics of the full ACCORD cohort and the analytic study cohort are shown in **Table 1**.

**Table 1.** Description of sample population.

|  | ACCORD Cohort | Study Cohort |
| --- | --- | --- |
| Patients, n | 10,251 | 6,853 |
| Age, years, mean (SD) | 62.8 (6.6) | 62.7 (6.6) |
| Female sex, n (%) | 3,952 (38.6 %) | 2,649 (38.7%) |
| Race and ethnicity |  |  |
| White, n (%) | 6,393 (62.4%) | 4,306 (62.8 %) |
| Black, n (%) | 1,953 (19.1%) | 1,280 (18.7 %) |
| Hispanic/Latino, n (%) | 737 (7.2%) | 453 (6.6 %) |
| Other, n (%) | 1,168 (11.4%) | 814 (11.9 %) |
| Clinical history and kidney status |  |  |
| History of cardiovascular disease, n (%) | 3,609 (35.2%) | 2,348 (34.3 %) |
| Diabetes duration, years, mean (SD) | 10.7 (7.6) | 10.6 (7.5) |
| Baseline eGFR, mL/min/1.73 m<sup>2</sup>, mean (SD) | 91.0 (27.2) | 91.5 (28.5) |
| Baseline UACR, mg/g, mean (SD) | 99.3 (362.4) | 87.1 (282.9) |
| Glycemic and cardiometabolic profile |  |  |
| HbA1c, %, mean (SD) | 8.1 (1.1) | 8.3 (1.0) |
| Fasting plasma glucose, mg/dL, mean (SD) | 175.2 (56.2) | 175.3 (55.4) |
| Systolic BP, mmHg, mean (SD) | 136.4 (17.1) | 136.8 (16.7) |
| LDL cholesterol, mg/dL, mean (SD) | 104.9 (33.9) | 106.2 (33.5) |
| HDL cholesterol, mg/dL, mean (SD) | 41.9 (11.6) | 41.9 (11.77) |
| Triglycerides, mg/dL, mean (SD) | 190.1 (148.4) | 189.3 (146.0) |
| BMI, kg/m<sup>2</sup>, mean (SD) | 32.2 (5.4) | 32.1 (5.4) |
| Medication use |  |  |
| ACE inhibitor or ARB use, n (%) | 7,102 (69.5%) | 4,752 (69.5 %) |
| Statin use, n (%) | 6,500 (63.7%) | 4,328 (63.4 %) |
| Metformin/biguanide use, n (%) | 6,554 (63.9%) | 4,372 (63.8 %) |
| Insulin use, n (%) | 3,582 (34.9%) | 2,379 (34.7%) |
| ACCORD trial assignments |  |  |
| Intensive glycemia assignment, n (%) | 5,128 (50.0%) | 3,394 (49.5%) |
| BP assignment: intensive BP, n (%) | 2,362 (23.0%) | 1,625 (23.7%) |
| Lipid assignment: fenofibrate, n (%) | 2,765 (27.0%) | 1,788 (26.1%) |

Baseline characteristics are shown for all randomized ACCORD participants and for the primary analytic cohort with baseline and 48-month eGFR available. Values are mean (SD) or n (%). Abbreviations: BP, blood pressure; eGFR, estimated glomerular filtration rate; UACR, urine albumin-to-creatinine ratio; ACE, angiotensin-converting enzyme; ARB, angiotensin receptor blocker; HDL, high-density lipoprotein; LDL, low-density lipoprotein; BMI, body mass index.

### Preprocessing

#### Baseline data

Baseline predictors were assembled from ACCORD analysis datasets and baseline case-report forms using the masked participant identifier. Data sources included the ACCORD key file, baseline laboratory files, blood pressure measurements, lipid measurements, concomitant medication records, and baseline history/physical examination data. Baseline eGFR, fasting plasma glucose, potassium and urine albumin-to-creatinine ratio (UACR) were extracted from the baseline laboratory file. HbA1c (glycated hemoglobin), blood pressure, and lipid variables were extracted from their respective baseline files. Baseline medication classes were converted to binary indicators. Baseline body mass index (BMI) was derived from measured weight and height.

Randomized treatment information was encoded using three binary indicators for assignment to intensive glycemia treatment, intensive blood-pressure treatment, and fenofibrate. For the blood-pressure and lipid indicators, participants assigned to standard/placebo and those not enrolled in the corresponding subtrial were coded 0.

The primary feature set included 98 baseline predictors before one-hot expansion: 25 continuous numeric variables, 59 binary indicators, and 14 categorical variables. Predictors spanned demographics and site/network, randomized treatment assignment, baseline kidney function, cardiometabolic laboratory values, vital signs, adiposity, medication classes, and medical history. Serum creatinine was excluded from the primary model because eGFR is derived from serum creatinine. Baseline urine biomarkers were also excluded from the primary urine-free model. UACR was retained only for clinical cohort description and sensitivity analyses.

#### Outcome definition

The primary outcome was annualized change in eGFR from baseline to 48 months, defined as:

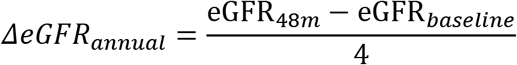

Where negative values indicate kidney function decline. Rapid decline was defined as:

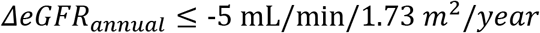

### Model Development

#### Model selection and training

We developed supervised regression models to predict annualized 48-month eGFR change, using baseline clinical variables only. Participants were split into training, calibration, and held-out test sets in a 60%/20%/20% ratio using fixed random seeds and stratification by outcome quantiles to preserve the distribution of kidney function change across splits. The training set was used for model fitting, the calibration set was reserved for conformal interval estimation, and the test set was used for final performance evaluation.

We compared several model classes with different assumptions about predictor-outcome relationships. Linear penalized models included ridge regression, lasso regression, and elastic net regression [18–20]. These models provide regularized linear benchmarks and reduce overfitting in the setting of many correlated baseline predictors. Tree-based models included random forest, histogram gradient boosting, and XGBoost [21–23], which can capture nonlinear associations and interactions among baseline variables. A mean-outcome dummy model, which predicted the training-set mean outcome for all participants, was included as a benchmark to quantify the added value of baseline clinical predictors.

All preprocessing was embedded within the model pipeline. Continuous variables were imputed using median imputation and then standardized. Categorical variables were imputed using most frequent category and then one-hot encoded. Binary variables were retained as 0/1 indicators with median imputation applied when missing. Preprocessing steps were fit only on the training data and then applied to calibration and test data to avoid information leakage.

Model performance was evaluated in the held-out test set using R^2^ and mean absolute error (MAE). MAE was emphasized because it is directly interpretable as the average absolute error in predicted annualized eGFR change, measured in mL/min/1.73 m². We also calculated relative MAE reduction compared with the dummy model:

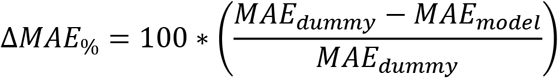

The best-performing primary model was selected by the lowest five-fold out-of-fold MAE within the training set. The calibration set was used only for conformal interval estimation, and the held-out test set was used only for final evaluation. Ninety-five percent confidence intervals for selected-model performance were estimated using 2,000 participant-level bootstrap resamples of the test set.

### Uncertainty quantification

Conformal prediction was used to convert point predictions into prediction intervals for individual future outcomes. For a nominal 90% interval, the target is that approximately 90% of future observed eGFR changes fall inside the interval. Empirical coverage was calculated in the held-out test set as the proportion of participants whose observed annualized eGFR change fell between the lower and upper interval bounds. Interval width was calculated as upper bound minus lower bound.

For the split conformal analysis, the trained point model was applied to the calibration set. For each calibration participant, we calculated the absolute residual as:

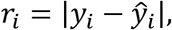

where *y_i_* is the observed annualized eGFR change and ŷ*_i_* is the predicted value. The conformal quantile of the calibration residual, *q*_0.90_, was then used as a common half-width for all test-set participants. Thus, the 90% split interval for a new participant was:

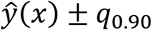

This method is simple and provides a global uncertainty margin, but it does not distinguish patients whose outcomes are easier or harder to predict.

For locally adaptive conformal prediction, we first estimated patient-specific residual scale. Within the training set, five-fold cross-validation produced out-of-fold point predictions. The corresponding absolute residuals were then used as the target for a residual-scale model, σ^(*x*), trained on the same baseline features. In the calibration set, normalized conformal scores were calculated as:

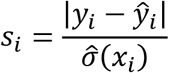

For a 90% interval, the conformal quantile of these normalized scores, *q*_0.90_, was used to construct locally adaptive intervals in the held-out test set:

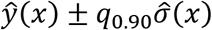

Therefore, two participants with the same predicted decline could receive different interval widths if one had a baseline profile associated with larger residual variability.

The locally adaptive interval width was treated as a patient-level reliability measure. Narrow intervals indicate that the model expects lower residual variability for that patient’s baseline profile, while wide intervals indicate that the prediction is less precise. Because the conformal quantile calibrates the residual-scale model, the intervals remain calibrated marginally even if the residual model is imperfect, provided the exchangeability assumption is reasonable.

### Predictability phenotype

The main uncertainty phenotype was the width of the 90% locally adaptive conformal interval. Held-out test participants were grouped into quintiles of interval width, from Q1 (narrowest intervals, most predictable) to Q5 (widest intervals, least predictable). We compared observed prediction error, rapid-decline prevalence, mean predicted and observed eGFR change, and baseline predictors across quintiles. We then used two complementary descriptive analyses: Spearman correlations between baseline variables and interval width, and logistic regression for membership in Q5.

Because baseline eGFR is a component of the change-score outcome, a prespecified robustness analysis reassigned width quintiles within 20 baseline-eGFR strata before pooling participants was performed. Associations of conditioned width with absolute prediction error and rapid decline were then adjusted for baseline eGFR.

### Interpretability

Model interpretability analyses were performed using SHapley Additive exPlanations (SHAP) [24] to characterize how baseline variables contributed to predicted 48-month eGFR change. SHAP values decompose each individual prediction into additive feature contributions relative to the model’s expected prediction. In this framework, each feature receives a contribution value for each participant. The magnitude of the SHAP value reflects how strongly that feature influenced the prediction, while the sign indicates whether the feature shifted the predicted annualized eGFR change toward a more favorable or more unfavorable kidney trajectory.

For the primary random forest model, SHAP values were calculated in the held-out test set. Global feature importance was summarized using mean absolute SHAP values, which quantify the average strength of each feature’s contribution across participants regardless of direction. This allowed us to identify the baseline variables most influential for point prediction of annualized 48-month eGFR change.

To connect interpretability with uncertainty, SHAP analyses were also summarized across quintiles of 90% locally adaptive conformal interval width. Participants were grouped from Q1, representing the narrowest intervals and most predictable predictions, to Q5, representing the widest intervals and least predictable predictions. Within each quintile, we calculated mean absolute SHAP values to determine which predictors contributed most strongly to point predictions among patients with low versus high prediction uncertainty. We then compared feature-importance patterns across quintiles to assess whether the predictors driving expected eGFR change differed between narrow-interval and wide-interval patients.

## Results

### Baseline Characteristics of the Study Population

Among 10,251 randomized ACCORD participants, 6,853 had baseline and 48-month eGFR measurements available and were included in the primary analytic cohort. The analytic cohort was broadly similar to the overall ACCORD cohort across baseline demographics, cardiometabolic risk factors, kidney function, and medication use (**Table 1**). Mean age was 62.7 years, 38.7% were female, and the racial/ethnic distribution was 62.8% White, 18.7% Black, 6.6% Hispanic/Latino, and 11.9% other race/ethnicity. Baseline cardiovascular disease was present in 34.3% of participants.

The study cohort had substantial cardiometabolic risk burden, with mean HbA1c 8.3%, mean systolic blood pressure 136.8 mmHg, mean BMI 32.1 kg/m², and mean triglycerides 189.3 mg/dL. Baseline kidney function was generally preserved, with mean eGFR 91.5 mL/min/1.73 m² and mean UACR 87.1 mg/g. Baseline medication use was common, including angiotensin-converting enzyme (ACE) inhibitor or angiotensin receptor blocker (ARB) use in 69.5%, statin use in 63.4%, metformin/biguanide use in 63.8%, and insulin use in 34.7%.

### Prediction of annualized eGFR change

The primary analysis evaluated prediction of annualized 48-month eGFR change using baseline clinical variables, excluding serum creatinine and urine biomarkers. Random forest had the lowest training-set cross-validated MAE (3.340 mL/min/1.73m²) and was selected before test-set evaluation. In the held-out test set, random forest achieved R^2^=0.382, MAE=3.271 mL/min/1.73m² (**Table 2**). Compared with the dummy mean-outcome benchmark, the random forest reduced MAE by 17.6%, demonstrating that baseline clinical features contained meaningful prognostic signal beyond the cohort-average trajectory.

**Table 2.**
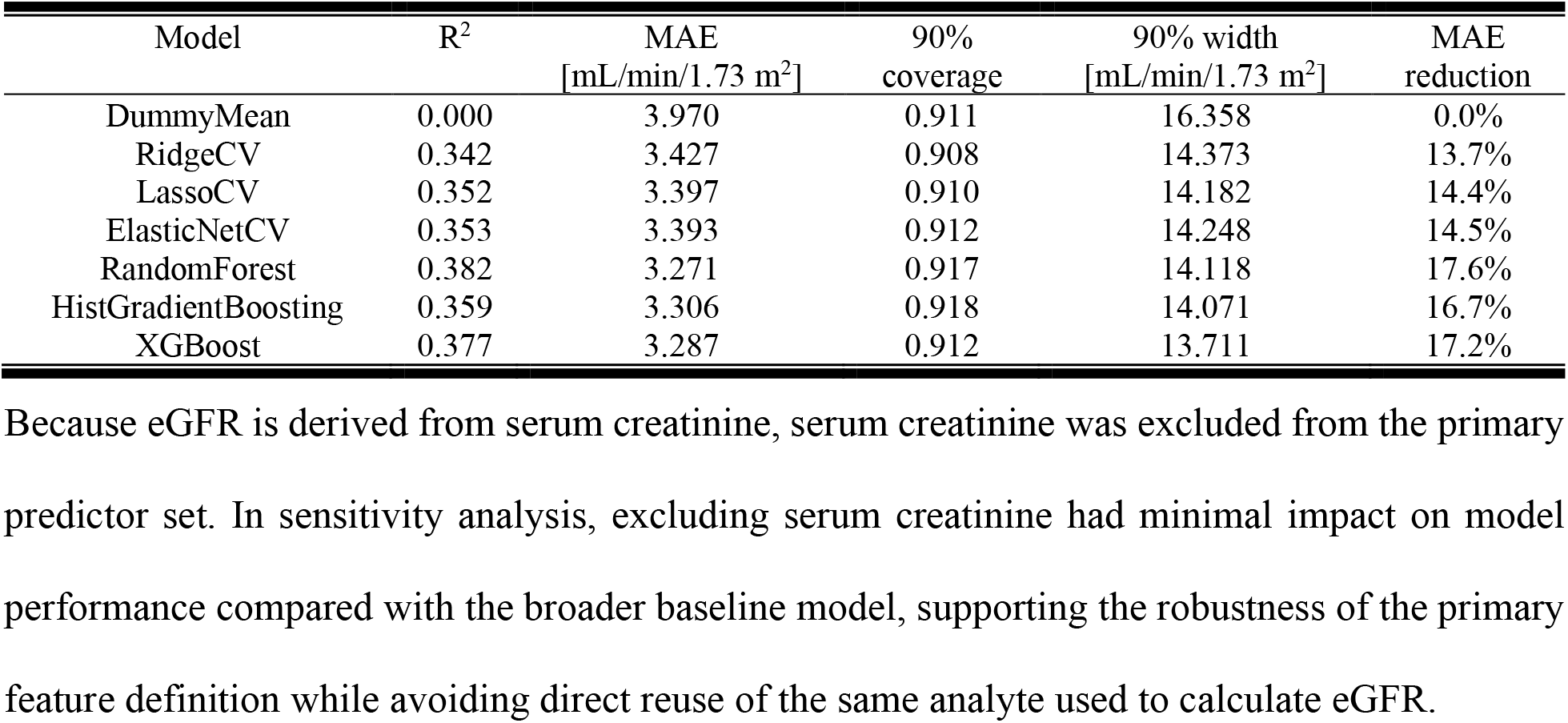
Held-out test-set performance for annualized 48-month eGFR change.

### Interpretability

In SHAP analysis of the best-performing random forest model, baseline eGFR was the dominant contributor to predicted 48-month annualized eGFR change, accounting for 36.0% of total mean absolute SHAP importance. Randomized fenofibrate assignment was the second most influential feature, followed by systolic blood pressure, intensive blood pressure assignment, triglycerides, age, Michigan Neuropathy Screening Instrument (MNSI) neuropathy score, alanine aminotransferase (ALT), HbA1c, and diabetes duration (**Figure 1**). Together, these results indicate that the model’s predictions were driven primarily by baseline kidney function, but also incorporated treatment assignment, blood pressure physiology, lipid-related markers, and diabetes severity. This pattern supports the clinical coherence of the model and suggests that long-term eGFR decline in this cohort was predictable from a compact set of baseline renal, cardiometabolic, and treatment-related characteristics.

**Figure 1.**
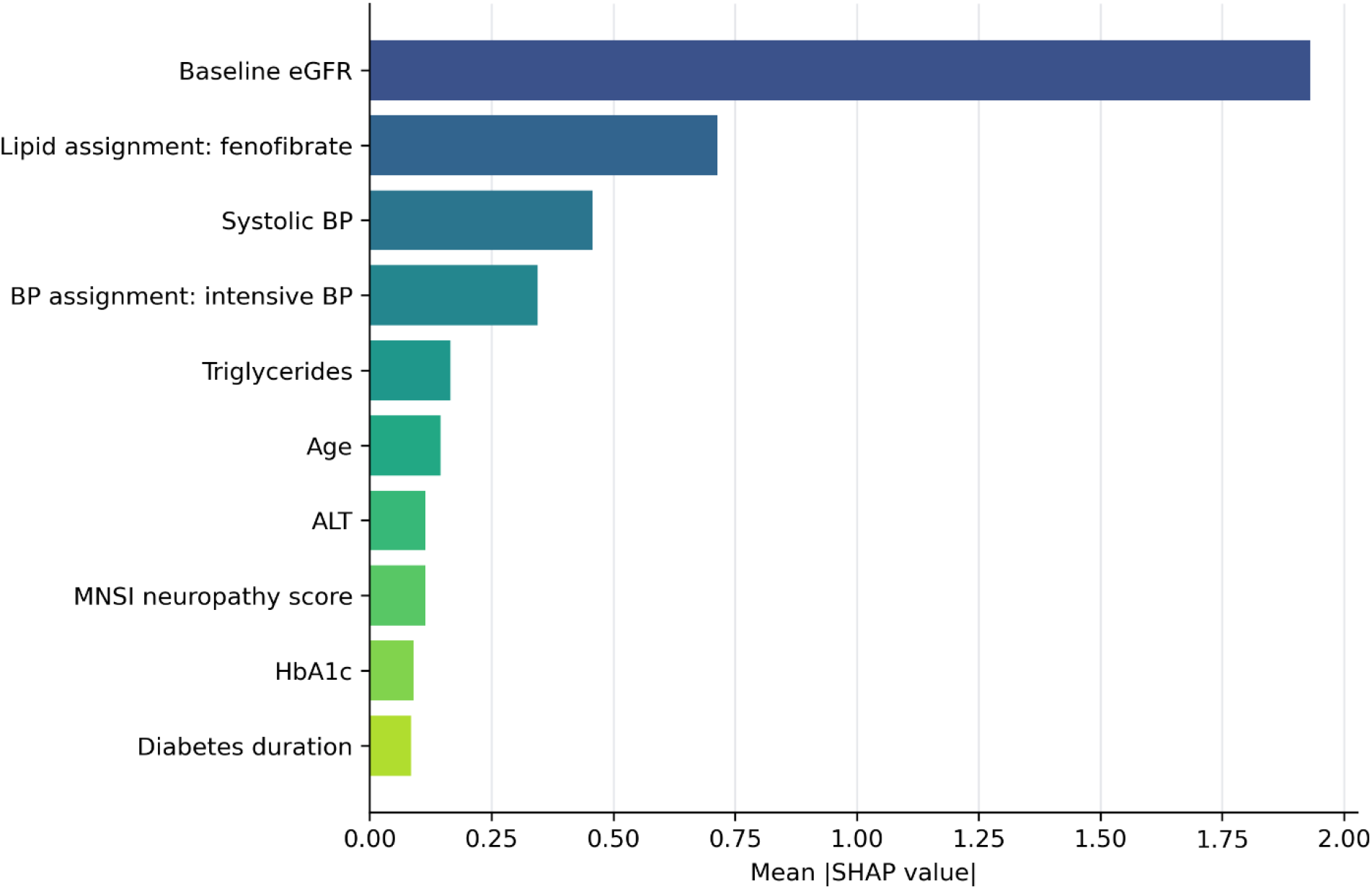
Model interpretability using SHAP values. Mean absolute SHAP values are shown for the 10 most influential predictors in the best-performing random forest model for 48-month annualized eGFR change. Larger values indicate greater average contribution to model predictions in the held-out test set.

### Conformal Calibration

Split 90% conformal intervals around the random forest predictions achieved empirical coverage of 0.917 with mean width 14.118 mL/min/1.73 m^2^ (**Table 2**). These intervals used one global residual half-width for all participants. Locally adaptive intervals were also well calibrated: empirical coverage was 0.795, 0.909, and 0.964 at nominal 80%, 90%, and 95% coverage, respectively. The locally adaptive 90% intervals had mean width of 13.759. Unlike split intervals, locally adaptive intervals used a patient-specific residual scale. Their width therefore became an analyzable patient-level quantity. Coverage was generally close to the 0.90 target across predicted-value quintiles, although some quintiles showed mild undercoverage or overcoverage consistent with finite subgroup size (**Figure 2**).

**Figure 2.**
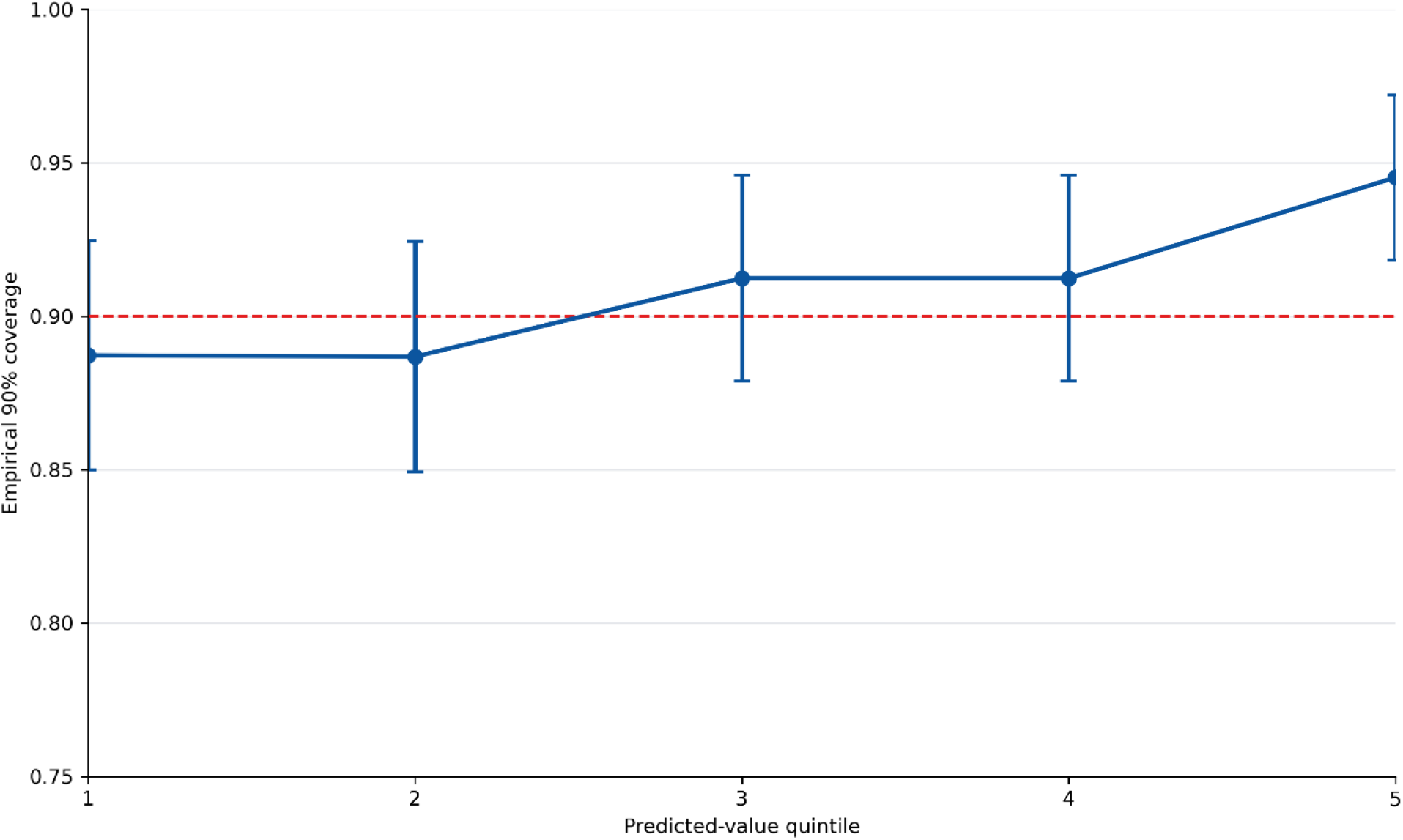
Coverage by predicted quintile. Empirical 90% locally adaptive conformal coverage by quintile of predicted annualized eGFR change. The dashed line indicates nominal 90% coverage; vertical bars are approximate binomial 95% confidence intervals.

### Predictability phenotype

In the initial unadjusted analysis, interval width identified patients whose outcomes were harder to predict and clinically more concerning. Mean 90% interval width increased from 12.11 mL/min/1.73 m^2^ in Q1 to 17.80 in Q5. MAE increased from 2.64 to 4.45, and rapid decline increased from 21.1% to 57.7% (**Figure 3**). Because baseline eGFR was strongly associated with width and is embedded in the change-score outcome, these unadjusted gradients were examined further in baseline-eGFR-conditioned analyses.

**Figure 3.**
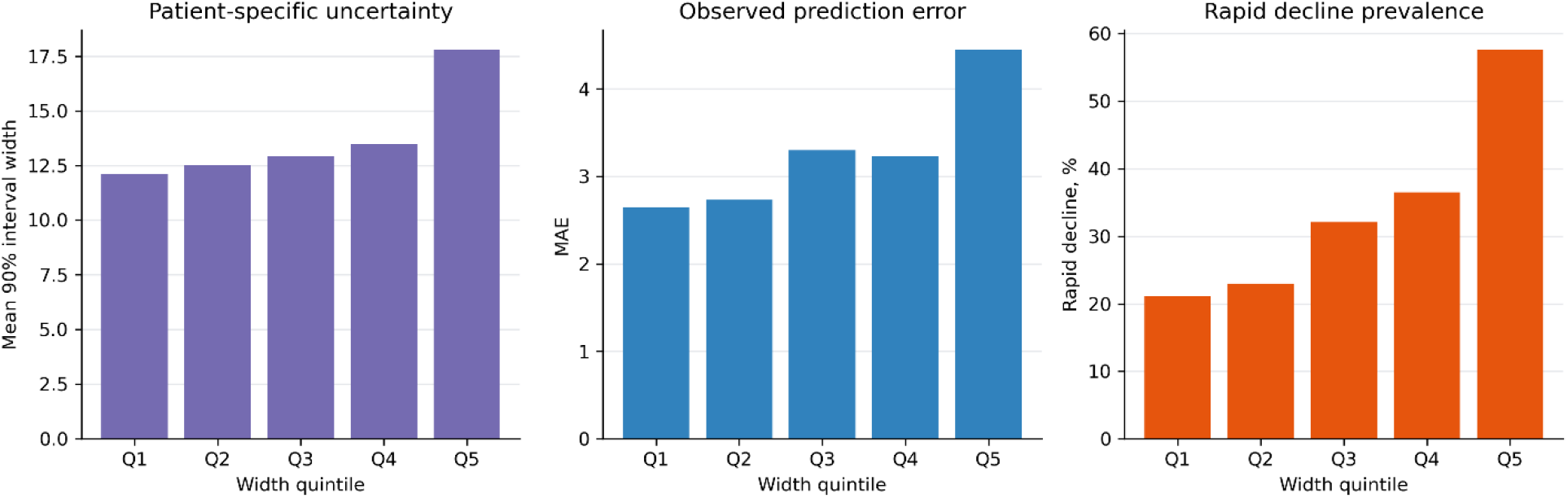
Unadjusted predictability phenotype. Participants were grouped by 90% conformal interval-width quintile. Wider intervals were associated with higher observed prediction error and higher rapid-decline prevalence.

The descriptive Q1-versus-Q5 table showed that participants with the widest intervals had substantially higher baseline eGFR, were younger, had higher HbA1c, included more women, and had higher triglycerides and systolic blood pressure (**Table 3**). Baseline eGFR was the dominant monotonic correlate of interval width, followed by age, HbA1c,female sex and triglycerides. These findings are descriptive and summarize how interval width varies across the held-out test set.

**Table 3.** Descriptive held-out test-set comparison of Q1 and Q5 interval-width groups.

| Feature | Q1 | Q5 | Spearman* |
| --- | --- | --- | --- |
| Baseline eGFR | 69.1 | 125.2 | 0.82 |
| Age | 69.0 | 59.7 | -0.454 |
| HbA1c | 8.1 | 8.9 | 0.207 |
| Female sex | 37.8% | 45.6% | 0.01 |
| Triglycerides | 165.8 | 224.0 | 0.08 |

\*Spearman is the rank correlation with 90% locally adaptive conformal interval width.

In the logistic model for membership in the widest-interval quintile, higher baseline eGFR, younger age, female sex, higher HbA1c, higher triglycerides and higher systolic BP were associated with Q5 membership (**Table 4**). The multivariable model distinguished participants in the widest interval-width quintile from those in Q1–Q4 with an apparent AUC of 0.945, supporting the idea that patient-level predictability is partly structured by baseline phenotype.

**Table 4.** Logistic regression for Q5 interval-width membership results, unadjusted for Baseline eGFR.

| Covariate | Scale | OR* | 95% CI | P |
| --- | --- | --- | --- | --- |
| Baseline eGFR | +10 mL/min/1.73 m<sup>2</sup> | 3.81 | 3.18-4.55 | <0.001 |
| Age | +5 years | 0.84 | 0.71-1.00 | 0.053 |
| Female sex | yes vs no | 1.78 | 1.15-2.78 | 0.01 |
| HbA1c | +1% | 1.96 | 1.61-2.38 | <0.001 |
| Triglycerides | +50 mg/dL | 1.08 | 1.00-1.17 | 0.064 |
| Systolic BP | +10 mmHg | 1.21 | 1.06-1.38 | 0.004 |

\*ORs above 1 indicate higher odds of widest-interval status.

These estimates were unadjusted for the strong relationship between baseline eGFR and interval width.

### Sensitivity analysis addressing baseline eGFR

After interval-width quintiles were assigned within 20 baseline-eGFR strata, the correlation between quintile and baseline eGFR was effectively removed (Spearman rho=0.00, P=0.782). MAE still increased from 2.935 in Q1 to 3.664 in Q5. Each one-quintile increase in conditioned width was associated with annual 0.151 mL/min/1.73 m^2^ greater absolute error after adjustment for baseline eGFR (95% CI, 0.049-0.253, P=0.004). By contrast, rapid decline was present in 37.9% of Q1 and 32.9% of Q5, and the adjusted association was not significant (OR per quintile, 0.96; 95% CI, 0.88-1.04, P=0.316) (**Figure 4**).

**Figure 4.**
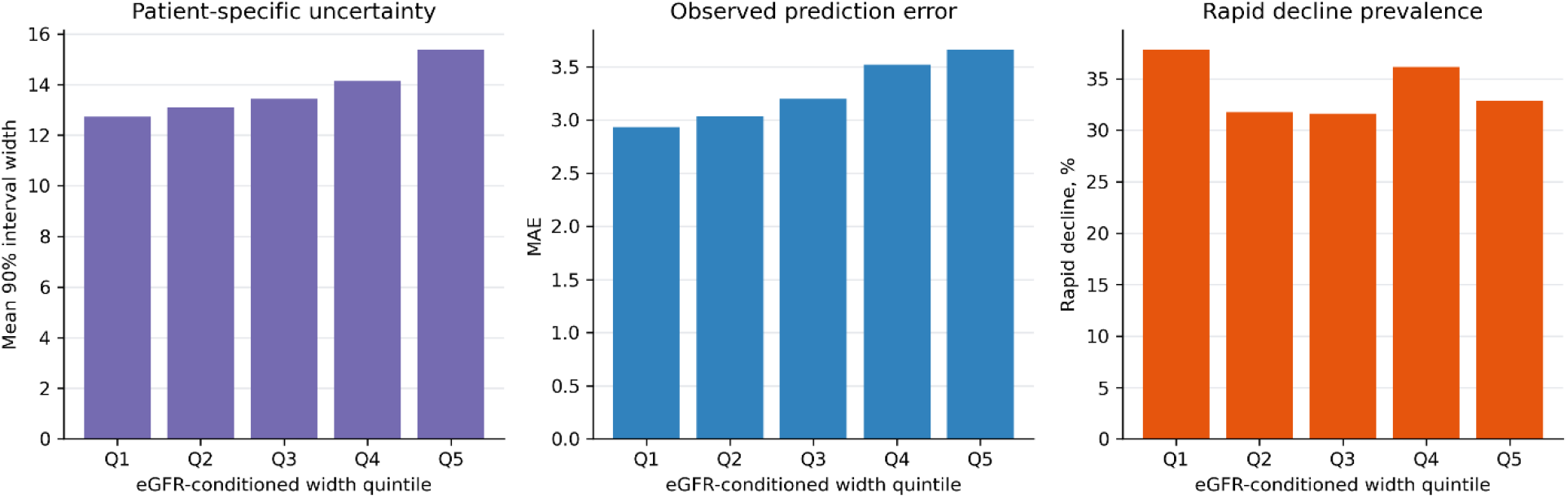
Adjusted predictability phenotype. Participants were grouped by 90% conformal interval-width quintile. Wider intervals were associated with higher observed prediction error, whereas rapid-decline prevalence showed no consistent gradient.

Importantly, conditioned interval width remained structured by the broader baseline phenotype. In a multivariable model for membership in the widest conditioned-width quintile, younger age, higher HbA1c, higher triglycerides, and higher systolic blood pressure remained associated with Q5 membership. Female sex remained directionally associated but was not statistically significant after conditioning (**Table 5**).

**Table 5.** Logistic regression for Q5 interval-width membership results, adjusted for Baseline eGFR.

| Covariate | Scale | OR* | 95% CI | P |
| --- | --- | --- | --- | --- |
| Age | +5 years | 0.68 | 0.60-0.77 | <0.001 |
| Female sex | yes vs no | 1.31 | 0.97-1.81 | 0.084 |
| HbA1c | +1% | 1.56 | 1.38-1.80 | <0.001 |
| Triglycerides | +50 mg/dL | 1.08 | 1.02-1.16 | 0.003 |
| Systolic BP | +10 mmHg | 1.14 | 1.04-1.26 | 0.003 |

\*ORs above 1 indicate higher odds of widest-interval status.

## Discussion

In this secondary analysis of ACCORD, baseline clinical variables predicted 48-month annualized eGFR decline with good long-horizon performance. The model improved meaningfully over the dummy mean predictor, demonstrating that baseline clinical state contains substantial information about future kidney trajectory. At the same time, the remaining error is clinically important and motivates uncertainty-aware decision support rather than point prediction alone.

The most novel finding is that prediction uncertainty itself was structured and clinically meaningful. In the unadjusted analysis, patients with the widest conformal intervals had higher observed prediction error and a much higher prevalence of rapid eGFR decline. However, the rapid-decline gradient was attenuated after accounting for baseline eGFR, consistent with mathematical coupling in the change-score outcome and regression to the mean. Interval width nonetheless retained an independent association with realized prediction error. Moreover, conditioned widest interval membership remained associated with younger age, higher HbA1c, higher triglycerides, and higher systolic blood pressure, indicating that prediction difficulty was structured by a broader baseline clinical phenotype rather than baseline eGFR alone. Thus, locally adaptive width is best interpreted as a calibrated, patient-specific reliability signal rather than an independent rapid-decline phenotype.

This framing fits the goals of medical informatics and decision-making. Clinical decision tools should ideally communicate both an estimate and the reliability of that estimate. A patient predicted to have steep decline with a wide interval may need closer surveillance, more frequent laboratory reassessment, or additional information gathering before high-consequence decisions. Conversely, a similar point prediction with a narrow interval may support greater confidence in the expected trajectory.

The decision to exclude serum creatinine was important methodologically because baseline and follow-up eGFR are derived from creatinine. Retaining baseline eGFR preserves clinically available kidney-function status, while avoiding direct reuse of serum creatinine as an additional predictor. The ACCORD-era eGFR equation also reflects older race-based kidney-function estimation, and should be revisited in external validation using contemporary race-free eGFR equations [25].

Fenofibrate assignment was the second-ranked contributor in the SHAP analysis. This finding should be interpreted in the context of fenofibrate’s known early and reversible effect on serum creatinine and, consequently, creatinine-derived eGFR [26,27]. In ACCORD, this increase reversed after treatment discontinuation and was not clearly indicative of progressive structural kidney injury. Therefore, part of the model’s reliance on fenofibrate assignment may reflect a treatment-related change in measured eGFR rather than underlying kidney disease progression.

Several limitations should be emphasized. First, this is internal validation within a clinical trial cohort, therefore external validation in contemporary diabetes populations is needed before clinical deployment. Second, ACCORD preceded widespread use of sodium-glucose cotransporter-2 (SGLT2) inhibitors and glucagon-like peptide-1 (GLP-1) receptor agonists, which may affect generalizability [28–31]. Third, conformal prediction provides marginal coverage under exchangeability, not guaranteed conditional coverage for every subgroup. Fourth, interval-width associations are descriptive and should not be interpreted as causal mechanisms for kidney decline.

## Conclusions

A baseline clinical model predicted 48-month eGFR decline in ACCORD with good long-horizon performance and calibrated conformal intervals. After accounting for baseline eGFR, patient-specific interval width remained associated with realized prediction error, and wider intervals remained associated with age, HbA1c, triglycerides, and systolic blood pressure. These results support uncertainty-aware kidney trajectory prediction as a clinically relevant direction for transparent decision support in T2DM populations.

## Data Availability

The deidentified ACCORD data analyzed during the current study are available through the National Heart, Lung, and Blood Institute BioLINCC repository, subject to repository application and data-use approval (https://biolincc.nhlbi.nih.gov/studies/accord/). Analysis code and derived aggregate outputs are available from the corresponding author on reasonable request.

## List of abbreviations

ACCORD: Action to Control Cardiovascular Risk in Diabetes
ACE: angiotensin-converting enzyme
ALT: alanine aminotransferase
ARB: angiotensin receptor blocker
BMI: body mass index
CKD: chronic kidney disease
eGFR: estimated glomerular filtration rate
GLP-1: glucagon-like peptide-1
HbA1c: glycated hemoglobin
MAE: mean absolute error
MNSI: Michigan Neuropathy Screening Instrument
OR: odds ratio
SGLT2: sodium-glucose cotransporter-2
SHAP: SHapley Additive exPlanations
T2DM: type 2 diabetes mellitus
UACR: urine albumin-to-creatinine ratio.

## Declarations

### Ethics approval and consent to participate

The Johns Hopkins Medicine Institutional Review Board of the Johns Hopkins University School of Medicine approved this secondary analysis (IRB00256340). The analysis used deidentified ACCORD data obtained through the National Heart, Lung, and Blood Institute BioLINCC repository. All participants provided written informed consent for participation in the parent ACCORD trial.

### Consent for publication

Not applicable.

## Authors’ contributions

D.O. led the study conception, analysis, and manuscript preparation. C.H. contributed to the methodology and analysis. P.S. and R.C. provided study oversight. C.P. contributed nephrology expertise and clinical interpretation. All authors critically reviewed and approved the final manuscript.

## Acknowledgements

Not applicable.

